# Strain inheritance and neonatal gut microbiota development: a meta-analysis

**DOI:** 10.1101/2020.10.10.20210534

**Authors:** Daniel Podlesny, W. Florian Fricke

## Abstract

As many inflammatory and metabolic disorders have been associated with structural deficits of the human gut microbiota, the principles and mechanisms that govern its initialization and development are of considerable scientific interest and clinical relevance. However, our current understanding of the developing gut microbiota dynamics remains incomplete. We carried out a large-scale, comprehensive meta-analysis of over 1900 available metagenomic shotgun samples from neonates, infants, adolescents, and their families, using our recently introduced SameStr program for strain-level microbiota profiling and the detection of microbial strain transfer and persistence. We found robust associations between gut microbiota composition and age, as well as delivery mode which was measurable for up to two years of life. C-section was associated with increased relative abundances of non-gut species and delayed transition from a predominantly oxygen-tolerant to intolerant microbial community. Unsupervised networks based on shared strain profiles generated family-specific clusters connecting infants, their siblings, parents and grandparents and, in one case, suggested strain transfer between neonates from the same hospital ward, but could also be used to identify potentially mislabeled metagenome samples. Following birth, larger quantities of strains were shared between vaginally born infants and their mothers compared to C-section infants, which further persisted throughout the first year of life and belonged to the same bacterial species as strains that were shared between adults and their parents. Irrespective of delivery type, older children shared strains with their mothers and fathers and, into adulthood, could be accurately distinguished from unrelated sample pairs. Prominent gut commensal bacteria were both among frequently transferred (e.g. *Bacteroides* and *Sutterella*) and newly acquired taxa (e.g. *Blautia, Faecalibacterium*, and *Ruminococcus*). Our meta-analysis presents a more detailed and comprehensive picture of the highly dynamic neonatal and infant gut microbiota development than previous studies and presents evidence for taxonomic and functional compositional differences early in life between infants born naturally or by C-section, which persist well into adolescence.

## INTRODUCTION

The taxonomic composition of the human microbiota is highly individual, with variations between individuals far exceeding those observed within one person over time (Costello et al., 2009). Although there is an ongoing controversy around prenatal microbiota exposure during pregnancy (Hornef and Penders, 2017), microbial colonization is generally believed to begin at birth. Birth type, i.e. vaginal delivery or Cesarean section (Blaser and Dominguez-Bello, 2016), and influences during the first weeks and months after birth, i.e. breastmilk or formula feeding (Bokulich et al., 2016), antibiotic or other medications (Cho and Blaser, 2012), exposure to family members (Korpela et al., 2018), rural or urban environments (Zuo et al., 2018), etc. have all been associated with compositional changes of the microbiota, resulting in the establishment of individualized and stable microbiota profiles by the age of 2-3 years (Cho and Blaser, 2012). Correspondingly, disruptive influences on early-life microbial exposure, such as C-section or microbiota depletion by antibiotic treatment, increase the risks for inflammatory and metabolic disorders (Mueller et al., 2015; Torow and Hornef, 2017).

Maternal microbiota transfer during vaginal birth has received considerable attention in the scientific literature (Stinson et al., 2018): Several highly cited studies reported an enrichment of the neonatal gut microbiota for skin and environmental bacteria in infants born by C-section as opposed to a taxonomic microbiota composition in naturally born infants, which resembled that of the vaginal microbiota (Dominguez-Bello et al., 2010; Reyman et al., 2019; Shin et al., 2015). Accordingly, retroactive colonization of the neonate with bacteria from the vaginal microbiota of the mother has been proposed to remediate potential microbiota deficits in infants born by C-section (Dominguez-Bello et al., 2016). However, this practice remains controversial, as it carries the risk of transferring (opportunistic) pathogens to the newborn and health benefits from this treatment have not been proven (Cunnington et al., 2016). Importantly, our understanding of the neonatal microbiota acquisition process has still many gaps, as the most commonly applied method for compositional microbiota analysis, 16S rRNA gene amplicon sequencing, does not provide sufficient resolution to track specific maternal or environmental strains in the neonatal microbiota. Recently, new bioinformatics tools have been applied for the differentiation of microbial subspecies lineages in the neonate, based on the detection of phylogenetic sequence variations in metagenomic shotgun sequence data (Ferretti et al., 2018; Korpela et al., 2018; Shao et al., 2019; Yassour et al., 2018). However, as these methods vary in sensitivity and specificity (Van Rossum et al., 2020) and most studies have only focused on the characterization of the early-life gut microbiota within the first year of life, the resulting findings remain difficult to compare and do not provide much insight into the long-term persistence of maternally transferred microbes in the infant, child and adolescent.

We sought to expand on previous studies by carrying out a large-scale, comprehensive meta-analysis of available metagenomic sequence data from neonates, infants and adolescents, their siblings, mothers and fathers and, in some cases, even grandparents. We used our recently introduced SameStr program (Podlesny and Fricke, 2020) for the sensitive, species-specific shared strains detection, in order to infer microbial transfer to the neonatal and infant microbiota, as well as microbial persistence during infancy, childhood and adolescence. We combined available metagenomes from nine distinct studies, comprising over 1944 samples, which represents the largest meta-analysis of the neonatal and infant microbiota so far. We report robust associations between gut microbiota composition and age beyond the first two years of life and compositional alterations in C-section infants, resulting in a delayed transition from a predominantly oxygen-tolerant to intolerant microbiota. We show that strain sharing during the first year of life was most prevalent in vaginally delivered infants and almost exclusively involved the intestinal microbiota of the mother, whereas older infants shared strains with both of their parents, irrespective of birth type. Strain transfer was taxonomy-dependent and involved several prominent gut commensals, such as members of the genera *Bacteroides, Bifidobacterium*, and *Parabacteroides*, although other gut bacteria (e.g. *Blautia, Faecalibacterium*, and *Ruminococcus*) were mostly represented by new, previously undetected strains.

## MATERIAL AND METHODS

### Metagenomic sequence data

Metagenomic shotgun sequences were collected from nine publicly available datasets, including samples obtained within the first week after birth (≤1W, n = 505), the neonatal period (≤1M, n = 202), early (≤6M, n = 241) and late (≤2Y, n = 170) infancy, childhood (≤10Y, n = 74), and samples taken from adolescents and adults (10Y+, n = 752), 625 of which were from collected from respective parents of the children (Table S1). For each subject, sequence data downloaded from the SRA were concatenated in case of multiple available accessions. Raw paired-end metagenomic shotgun sequence reads were quality processed with kneaddata (KneadData Development Team, 2017). Sequence regions where base quality fell below Q20 within a 4-nucleotide sliding window were trimmed and removed if they were truncated by more than 30% (SLIDINGWINDOW:4:20, MINLEN:70) or aligned with the human genome (GRCh37/hg19) with bowtie2 (Langmead, 2010). Output files consisting of surviving paired and orphan reads were concatenated and used for further processing.

### Taxonomic microbiota profiling and functional annotation

Preprocessed sequence reads from each sample were mapped against the clade-specific marker gene database using MetaPhlAn2 (Truong et al., 2015). Relative abundances of species-level taxonomic profiles were centered-log ratio (clr) transformed and used for principal components analysis (PCA, FactoMineR). For taxonomic analyses we further aggregated functional metadata on bacterial species (Table S2) from (Browne et al., 2016), (Vatanen et al., 2019), List of Prokaryotes according to their Aerotolerant or Obligate Anaerobic Metabolism (OXYTOL 1.3, Mediterranean institute of infection in Marseille), bacDive (Reimer et al., 2019), FusionDB (Zhu et al., 2017), The Microbe Directory v 1.0 (Shaaban et al., 2018), and the expanded Human Oral Microbiome Database (Escapa et al., 2018).

### Strain sharing across biological samples

Shared strains were identified with the recently released SameStr program (Podlesny and Fricke, 2020), leveraging within-species phylogenetic sequence variations in clade-specific marker genes. For this, MetaPhlAn2 marker alignments were converted to single nucleotide variant (SNV) profiles, filtered, merged and compared between metagenomes based on maximum variant profile similarity (MVS) that takes all detected single nucleotide variants (SNVs) into consideration, including polymorphic sites (≥10% allele frequency) resulting from multiple strains representing the same species. Shared strains are called for overlapping species alignments between metagenomic samples of ≥5kb length and with a MVS of ≥99.9%. The SameStr program and further documentation is available at github: https://www.github.com/danielpodlesny/SameStr.git

### Statistical Analyses and Visualizations

We used species and genus-level microbiota profiles to model the chronological age of the sample host. For this, we trained a Random Forest regression model (randomForest) on data from vaginally delivered infants and adults (80% data split) and evaluated the predictions (root mean square error (RMSE), caret) using data from the test set (20%) and C-section delivered infants. Variable importance was determined by quantification of mean increase of mean squared error (MSE) after permuting each variable. To assess the impact of delivery mode on taxonomic microbiota composition (Figure 2B) we calculated PERMANOVA (adonis, vegan) with 1e4 permutations for fecal samples collected within given time ranges. Related mother/child pairs were identified using simple logistic regression based on the number and fraction of shared strains between sample pairs. We divided data into training and test data (60% / 40%) and evaluated the classifiers on hold-out data of vaginally and sectio-delivered infants, including calculation of the precision-recall (tidymodels) and receiver operating characteristic (ROC) curves (plotROC) and their statistics. Microbiota composition was visualized across samples (Figure 2C, 5C) with area plots using binomial smoothing (glm, logit-link) of relative abundances. Strain co-occurrence detected with SameStr (Figure 4) was visualized in networks generated with tidygraph (stress layout), showing sample connections with more than four shared strains.

## RESULTS

### Maturation of the taxonomic gut microbiota composition is linked to age

In order to carry out a comprehensive characterization of the newborn microbiota, its development over the first weeks and months of life, and maturation during infancy and into adolescence and adulthood, we compiled a comprehensive dataset of over 1,700 fecal metagenomes from nine recent studies for meta-analysis (see Table 1 for overview). This dataset included samples from 441 infants from 415 different families, corresponding samples from 383 mothers and 16 fathers, as well as cross-sectionally collected samples from eleven individuals from two families of three generations. A complete list of all samples and metadata is included in the supplement (Table S1).

**Table 1:**
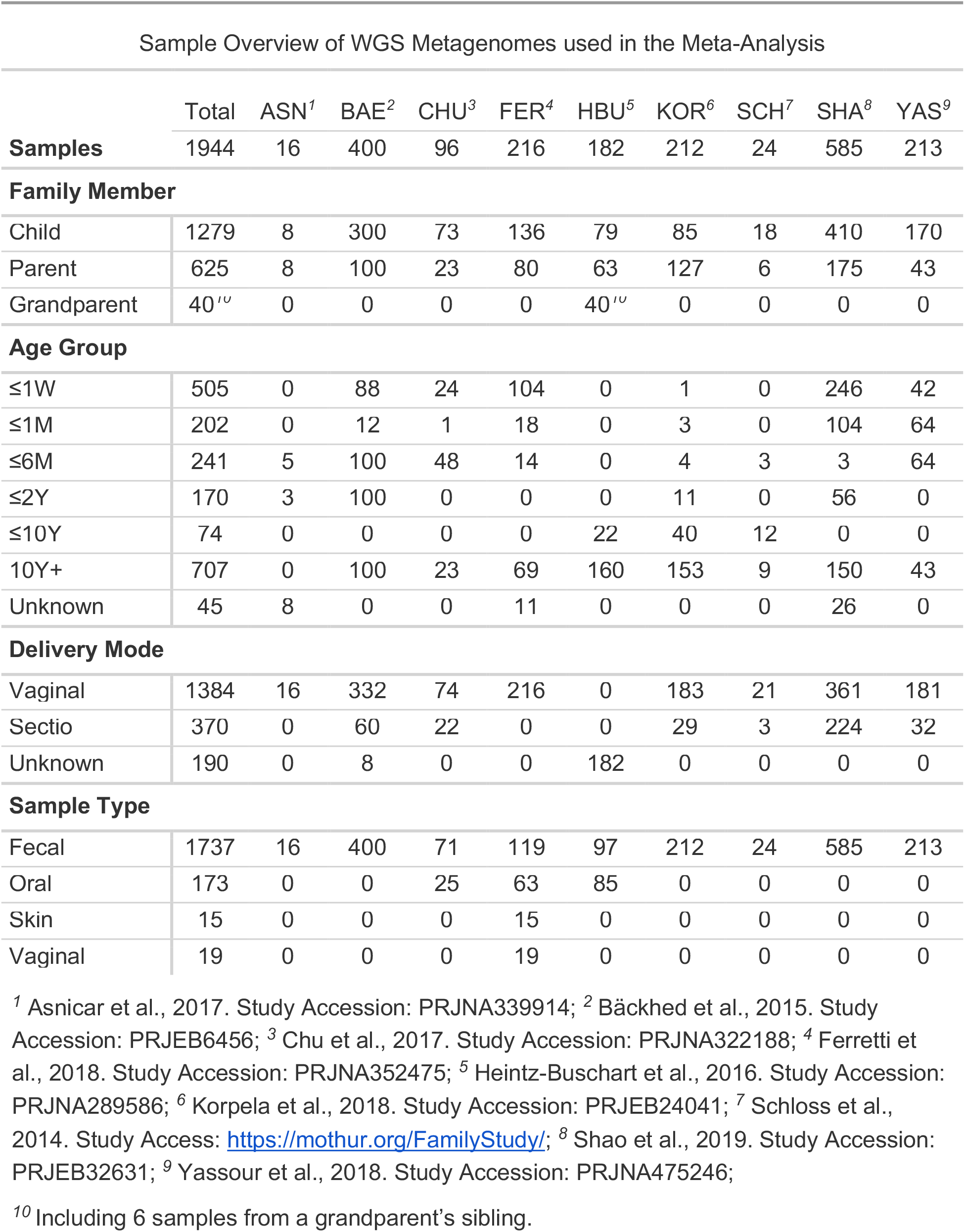
Overview of Metagenomic Sequencing Data used in the Meta-Analysis.

High-resolution taxonomic characterizations of all metagenomes were carried out with MetaPhlAn2 (Truong et al., 2015) and used to compare overall (beta-diversity) compositional profiles between samples at the genus and species-level. Taxonomic microbiota compositions could be linked to age, with adult and infant samples comprising distinct clusters in principal component analysis (PCA; Figure 1A, S1A). Within-sample alpha diversity served as a major distinguishing factor, which was strongly correlated with PC1 of the beta-diversity analysis (Figure 1B, spearman r=0.73, p<0.001 for Shannon index). Taxonomic microbiota compositions have previously been associated with chronological age, based on 16S rRNA amplicon sequence data (Galazzo et al., 2020; Subramanian et al., 2014) or metagenomic sequence data of limited resolution (only three samples per infant; (Bäckhed et al., 2015)). To determine if taxonomic microbiota compositions in our larger dataset were linked to age and to identify those microbial taxa with the strongest informative value for age prediction, a random forest regression model was trained on 80% of compositional microbiota profiles from vaginally delivered infants or adults (training dataset). The accuracy of the resulting model was evaluated on hold-out samples (test dataset) and demonstrated a strong positive correlation between predicted and actual age, including for adults (Figure 1D; RMSE=0.42; Figure S1B). The relative abundances of bacterial genera that include typical gut (e.g. *Vellonella, Bifidobacterium, Faecalibacterium*) but also skin (*Staphylococcus, Propionibacerium*) commensals were found to be most informative for the model (Figure 1C). Thus, the meta-analysis of our combined dataset confirms the association of taxonomic fecal microbiota composition with the chronological age of the human host, which extends beyond the first 2-3 years of life that have previously been suggested for the maturation into its adult composition (Arrieta et al., 2014).

**Fig. 1:**
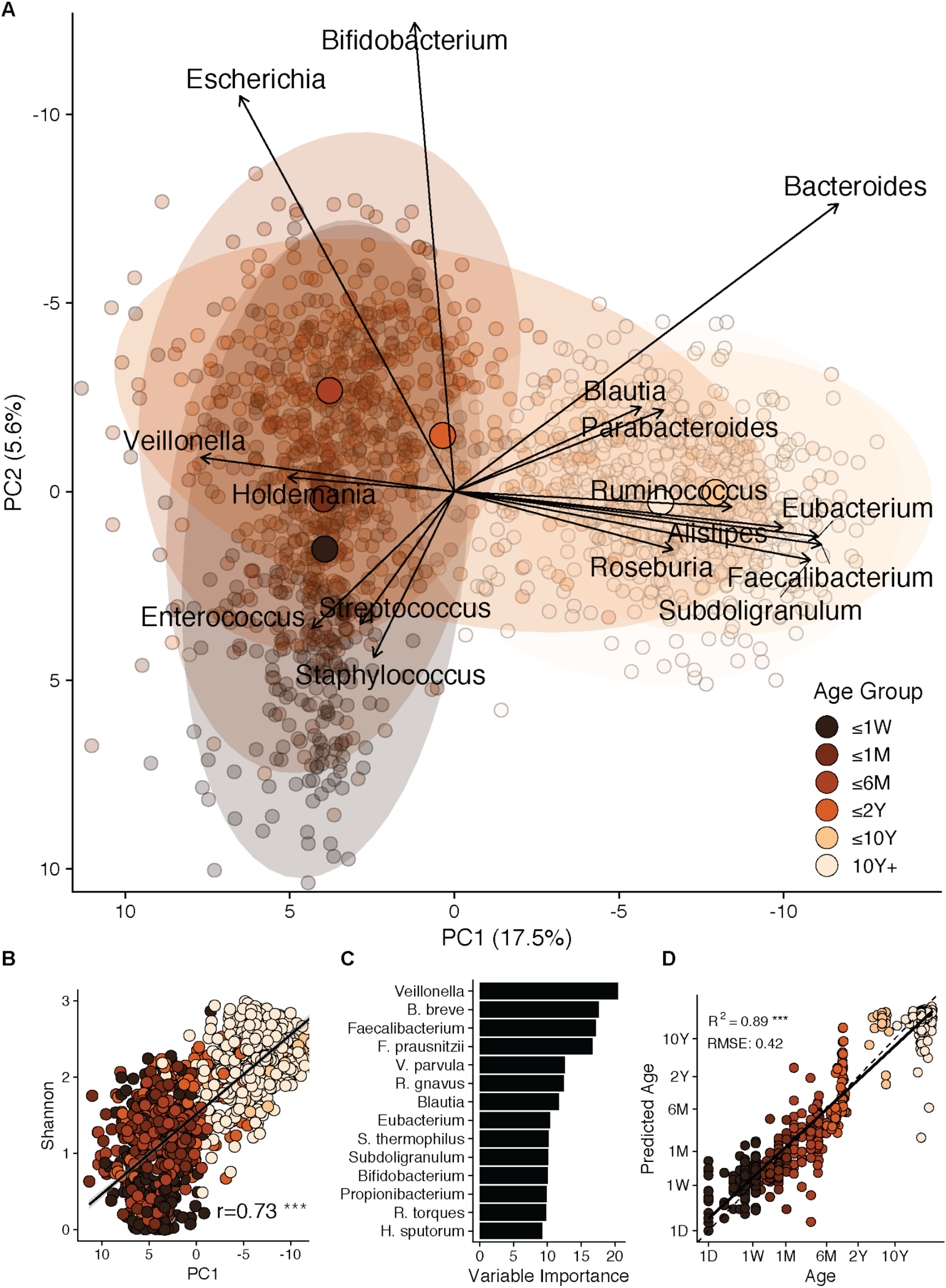
Maturation of the taxonomic human gut microbiota composition is linked to age. A, B): Principal component analysis (PCA) biplot of centered log-ratio (clr)-transformed genus-level (species-level in Figure S1A) microbial community compositions of 1,679 human fecal samples obtained from neonatal stage up to late adulthood (range: 0 - 72 years) shows a strong compositional clustering along principal components PC1 and PC2 (A), which is correlated with microbial alpha-diversity (B). By the age of 2 years the developing microbiota reaches the level of alpha-diversity seen in adults. C, D): Compositional changes during microbiota maturation are predictive of host age (D). Key taxa used by the random forest regression model include gut commensal genera, such as *Veillonella, Bifidobacterium*, and *Faecalibacterium*, as well as bacterial genera associated with the skin microbiota, such as *Staphylococcus* and *Propionibacterium* (C).

### Delayed transition from predominantly oxygen-tolerant to intolerant microbiota in C-section infants

As delivery mode has repeatedly been associated with compositional alterations of the neonatal microbiota (Bazanella et al., 2017; Shao et al., 2019; Stewart et al., 2018), we stratified our combined dataset for infant birth type by vaginal delivery or C-section. During the first week of life, birth type had a strong impact on microbiota compositions, explaining up to 4% of the taxonomic variation as determined by PERMANOVA (Figure 2A, p = 1e-6, adonis function, 1e4 permutations). However, measurable associations of delivery type and microbiota composition decreased over time, with only 1.3% of total taxonomic variation explained by C-section in 2-year-old infants (p = 0.05), which was comparable to the fraction of observed variations (1.1%, n.s.) that was associated with gender (Figure 2B). On the genus and species level, the largest differences between vaginally delivered and C-section infants were observed before six months of age and included increased relative abundances of the bifidobacteria *B. longum* and *B. bifidum*, the *Bacteroidales* species *B. dorei, B. fragilis, Parabacteroides distasonis* and the *Enterobacteriaceae* species *Escherichia coli* in vaginally delivered infants. C-section infants were characterized by increased relative abundances of species from the genera *Enterobacter, Klebsiella, Streptococcus*, the family *Veillonellaceae*, and *Enterococcus faecium* (Figure 2C), suggesting higher colonization rates of opportunistic pathogens as previously described (Shao et al., 2019).

**Fig. 2:**
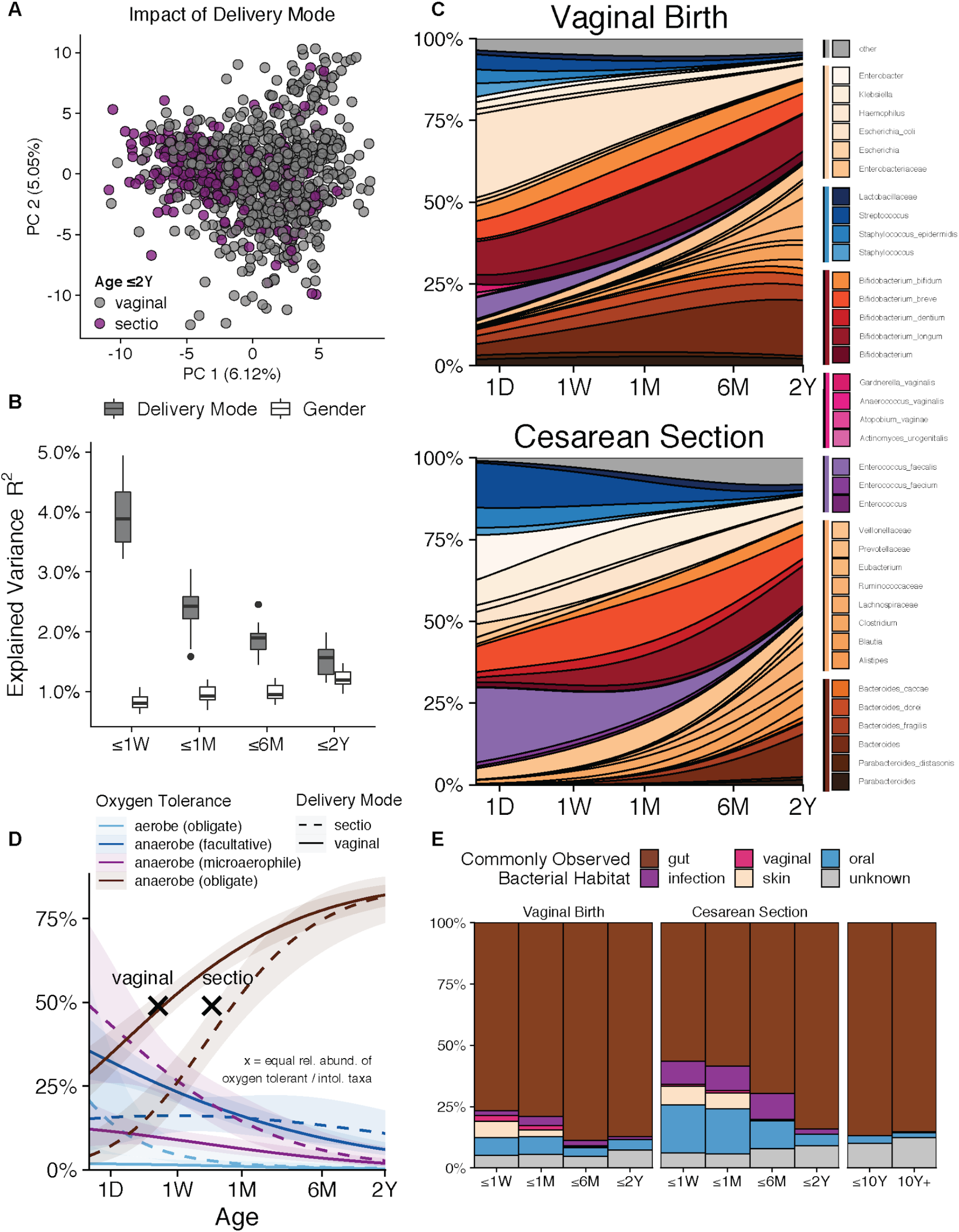
Delayed transition from predominantly oxygen-tolerant to intolerant microbiota in C-section infants. A) Differences in taxonomic gut microbiota compositions within the first two years after birth distinguish vaginally born and C-section infants. B) Delivery mode explains up to 4% of taxonomic variation during the first week of life, but the impact of delivery mode declines over time, as determined by PERMANOVA at different time points (adonis function, 1e4 permutations). C) Relative abundance of genus and species-level taxa modelled across 1032 fecal samples over time (glm, binomial smoothing), split by delivery mode. Infants delivered by C-section are depleted of the genera *Bacteroides* and *Parabacteroides*, but show increased relative abundance of *Enterococcus faecalis, Klebsiella*, streptococci and staphylococci. D) For the first few years of life, the gut microbiota of vaginally delivered infants harbors larger fractions of obligate and facultative anaerobe species and lower fractions of microaerophile and strict aerobe species compared to C-section infants. E) The gut microbiota of neonates and infants (<2 years) contains larger proportions of species that have a preferred oral, skin or vaginal habitat or are associated with infection. This predicted non-gut microbiota fraction is disproportionately increased in C-section infants.

To complement the taxonomic analysis with functional insights, available phenotype information was used to assign species to various functional categories, based on oxygen requirements and preferred habitat or growth context (Table S2). Total microbiota fractions assigned to oxygen-tolerant and intolerant species differed substantially between vaginally delivered and C-section infants (Figure 2D). In general and across all infants, microbiota fractions of oxygen-tolerant species continually decreased, whereas those of oxygen-intolerant species continually increased over the first two years of life. However, in vaginally delivered infants, oxygen-tolerant microbiota proportions were predominantly represented by facultative anaerobe species (mean cumulative relative abundance: 27.2% vs. 15.8% in sectio; e.g. *Escherichia coli, Staphylococcus epidermidis, Klebsiella pneumoniae*) but microaerophile species in C-section infants (mean cum. rel. abundance: 10.9% vs. 32.9% in sectio; e.g *Enterococcus faecalis, E. faecium, Streptococcus vestibularis*). Moreover, vaginally delivered infants started with larger proportions of strict anaerobe species (mean cum. rel. abundance: 48% vs. 33% in sectio; e.g. *Bacteroides fragilis, B. vulgatus, B. dorei*), which constituted >50% of the total microbiota by day 4. On average, this transition to a predominantly obligate anaerobe gut microbiota was delayed by 10 days in C-section infants. This delay is noteworthy, as microbiota maturation in general, based on the taxonomic alterations described above, showed no difference between vaginally born and C-section infants (Figure S1B). Thus, our findings are in agreement with recent reports of increased colonization with opportunistic pathogens in neonates born by C-section (Shao et al., 2019) and provide a functional extension of the suggested “stunted microbiota” phenotype as potentially associated with delayed microbiota transition to a predominantly oxygen-intolerant species composition.

Besides oxygen intolerance, the preferred species habitat was another distinguishing feature that separated vaginally delivered from C-section infants (Figure 2E). Only 2.6% ± 5 of the older infant (>10 years) and adult gut microbiota were ascribed to non-gut or oral species (e.g. *Streptococcus salivarius, Veillonella parvula*), but these species contributed 17% ± 25.6 and 35.7% ± 32.4 to the neonatal microbiota of vaginally delivered and C-section infants, respectively. Species of predicted vaginal origin (e.g. *Gardnerella vaginalis, Atopobium vaginae*, and the lactobacilli *L. crispatus, L. iners)* represented significant neonatal microbiota fractions only within the first days of life (2.5% ± 11.4 vs. 0.7% ± 5.4 in vaginally born and C-section infants) and species that are commonly associated with infections (e.g. *Klebsiella oxytoca, Citrobacter koseri*) were commonly found in the early infant (1 day to 6 months) microbiota, in particular in infants born by C-section (9.2% ± 20.5% compared to 2.3% ± 9 for vaginally delivered infants).

In summary, C-section was associated with altered gut microbiota compositions, delayed transition from a predominantly oxygen-tolerant to non-tolerant microbiota and increased relative abundance of atypical gut species, suggesting that disrupted microbiota initialization in C-section infants could make the neonatal ecosystem more permissive for the colonization with disadvantageous species and opportunistic pathogens.

### Shared strain profiles distinguish related and unrelated mother/infant pairs

Compositional deficits of the neonatal microbiota of infants born by C-section have been attributed to reduced maternal microbiota transfer (Shao et al., 2019). However, species or genus-level microbiota compositions provide insufficient taxonomic resolution to infer microbial transfer, as distinct sub-lineages within genera or even species can be shared between unrelated samples or individuals (Lloyd-Price et al., 2017). Therefore, in order to detect microbial transfer between infants and their parents and microbial persistence in infants over time, we used the recently developed SameStr program from our group (Podlesny and Fricke, 2020) to identify shared unique subspecies taxa, i.e. strains, between metagenomes from our combined dataset. Various definitions of microbial strains exist, based on biological and evolutionary concepts, as well as phenotypic or phylogenetic detection methods (Van Rossum et al., 2020; Yan et al., 2020). SameStr applies a conservative approach to call shared strains, using restrictive thresholds to detect highly similar subspecies genetic variants in distinct metagenomic samples, as indication for strain persistence or transfer between the corresponding microbiomes (Podlesny and Fricke, 2020). Shared strains would be unique in the sense that the corresponding subspecies lineages would only be found in related metagenomes (e.g. from infant/mother pairs or longitudinally collected samples from the same individual) but not in unrelated metagenomes (e.g. from distinct infants or mothers). To test the specificity of SameStr’s shared strain calls on our combined dataset, adult and infant (0 - 60 years) samples were divided into related (independent samples from the same individual) and unrelated (samples from independent individuals) pairs and compared in regard to shared strain predictions (Figure 3A). Both in adults and infants, shared strains were commonly detected in related (18.3 ± 12.4; 6.2 ± 8.3) but not unrelated sample pairs (0.3 ± 0.6; 0.1 ± 0.3), whereas unrelated sample pairs showed a high overlap in species and genus-level taxon profiles (Figure 3A). Thus, these findings attest to the sensitivity and low false positive rate of unique shared strain predictions with SameStr and the utility of shared strain profiles to infer microbial transfer or persistence.

**Fig. 3:**
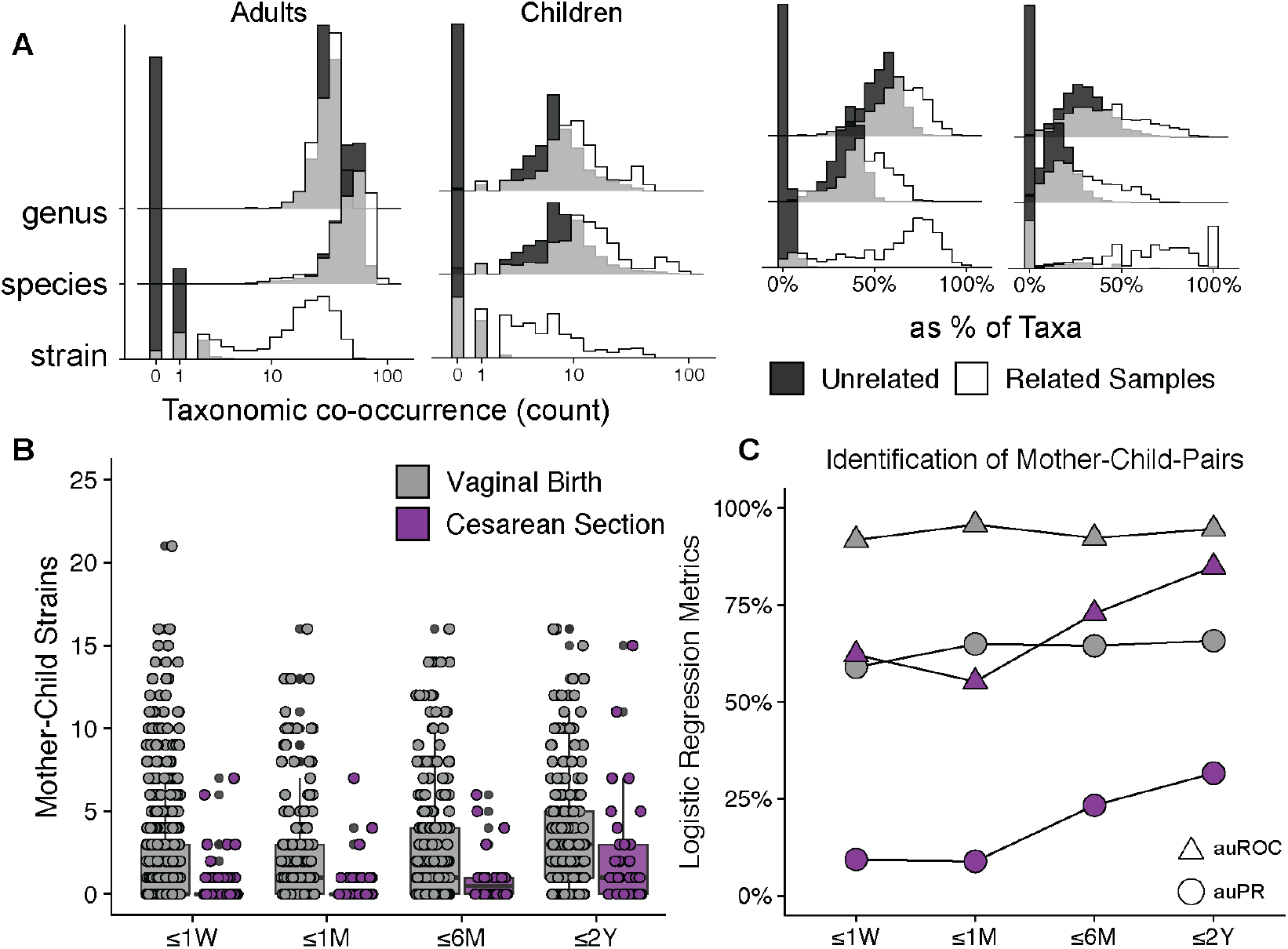
Shared strain profiles distinguish related and unrelated mother/infant sample pairs. A) Distributions of shared strains, species, and genera in >300.000 related and unrelated sample pairs, as total counts (left) and fractions of all detected taxa (right), highlight the specificity of SameStr’s shared strain calls. B) Counts of strains shared between vaginally and C-section delivered infants and their mothers within the first two years of life. C) Logistic regression models based on shared strain profiles can accurately identify mothers of vaginally but not C-section delivered neonates and infants (⪬2 years), as compared using area under the Receiver Operating Characteristics (auROC) and area under Precision/Recall (auPR) curve values.

To test if shared strain profiles between vaginally delivered and C-section infants during the first two years of life (Figure 3B) were sufficient to predict related infant/mother pairs, all sample pairs were divided into a training and test dataset (60/40 split) and shared strain profiles used as input for a random forest classifier (Figure 3C). For the prediction of vaginally born infant/mother pairs, the classifier achieved high and consistent accuracy (area under the Receiver Operating Characteristics (auROC) = 92-95%; area under Precision/Recall (auPR) = 59-66%; true positive (TP) rate = 1.42%), which was markedly reduced for C-section infant/mother pairs (auROC = 55-85%; auPR = 9-32%; TP rate = 0.93%), in particular during the first month of life but also at later time points.

### Construction of family networks based on shared strain calls

In order to further assess the extent and specificity of family-specific microbiota exchange, shared strain profiles were used to generate unsupervised networks showing individual samples as nodes and shared strain numbers as edges (Figure 4). Within these networks, samples were generally assigned to family-specific clusters, which in many cases included infants, separate siblings, mothers and fathers (Figure 4A), as well as children, parents and grandparents (Figure 4B), from the same but not distinct families.

**Fig. 4:**
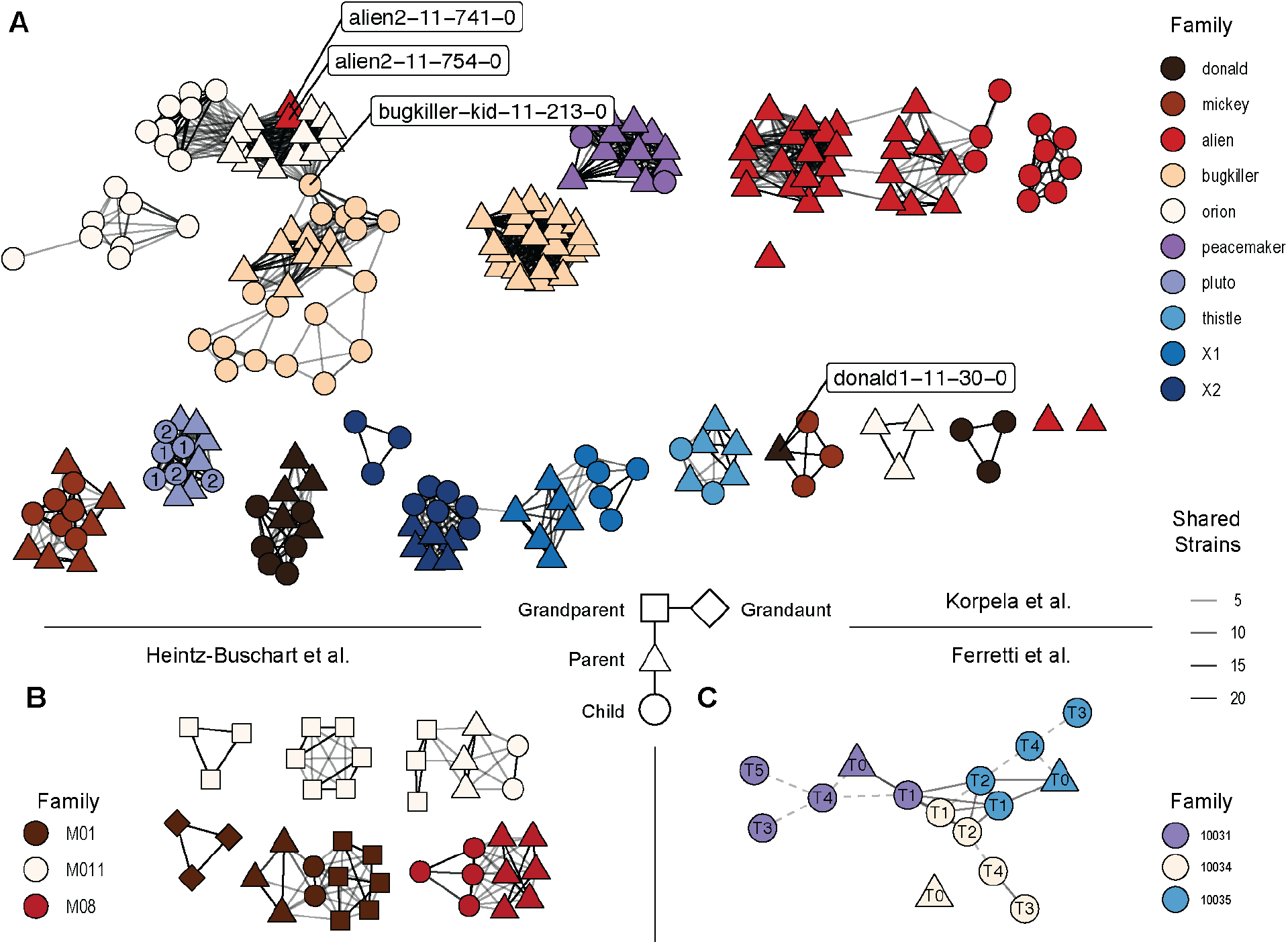
Shared strain networks identify microbiota exchange between multi-generational family members and distinct neonates. Network plots showing. A) family-specific strain sharing and potentially mislabeled or cross-contaminated samples; children of family “pluto” are exemplarily numbered, highlighting strain sharing between siblings B) multi-generational strain sharing involving grandparents; and C) strain sharing between distinct newborns from the same neonatal hospital ward, numbered according to sampled day after birth. Dashed lines show connections with 2-4 shared strains.

Network analysis also identified several inconsistent samples (Figure 4A). These samples, instead of sharing strains only with members from their own family, were connected to multiple samples from a different family (“donald1” and “alien2”) or to members from two different families (“bugkiller”). The inconsistent findings never included samples from different studies, but instead involved apparently random infant and parent samples and time points: in case of “alien2” another sample was collected from the same individual two days after “alien2-11-741-0” and 11 days before “alien2-11-754-0”, which shared no strains with these samples but with several samples from related family members. As there was no obvious biological explanation for these inconsistencies, which could result from human error (e.g. mis-labeling of the “alien2” or “donald1” samples) or technical problems (e.g. cross-contamination of the “bugkiller” sample), the conspicuous samples were removed from further analysis.

We observed one case in our combined dataset, which suggests that microbial strain exchange with non-family members could take place early-on during the neonatal microbiota development: Five fecal samples from three distinct neonates that were collected during the first two days of life shared strains with each other (Figure 4C), as well as with fecal, skin and vaginal samples from their mothers (Figure S2). No strain sharing was detected between samples collected at later time points (1 week - 6 months after birth). All three infants shared between 7 and 9 strains with each other, none of which belonged to typical intestinal commensals but bacterial species typically assigned to the oral, vaginal or skin microbiota (*Atopobium vaginae, Gardnerella vaginalis, Lactobacillus crispatus* and *L. iners, Prevotella buccalis* and *L. timonensis, Prevotella bivia*, and *Propionibacterium acnes*). The same species were also identified in other metagenomes from the combined dataset but never in other unrelated sample pairs. The infants were born at the same neonatal ward and possibly sampled at overlapping time points (Ferretti, personal communication). Thus, overlapping microbiota strain profiles could have resulted from microbiota exchange during the stay of infants and mothers in the clinic, e.g. through family members, nurses and other personnel, although cross-contamination during sample collection, storage and processing can also not be ruled out. In any case, these findings demonstrate the utility of subspecies taxonomic microbiota profiling to check public or new metagenomic sequence data for inconsistencies and improve our understanding of developmental microbiota processes involving microbial transfer and exchange.

### Maternal strain transfer in neonates and persistence through infancy and adolescence

To assess the role of microbiota transfer from family members to the developing infant gut microbiota, we first compared shared strain profiles in infants with different microbiome niches from the mother (Figure 5A), considering only cases from our combined dataset for which metagenomic sequence data from multiple maternal body sites were available (Ferretti et al., 2018; Heintz-Buschart et al., 2016). The largest fraction of the vaginally born infant microbiota was represented by shared strains with the intestinal microbiota of the mother, both immediately after birth (day 0-7: 30.6% ± 38.3 of the infant microbiota) and during the following weeks and months (day 30-120: 29.2% ± 26.5). Shared strain contributions from other maternal body sites were small (1.12% ± 7.76 relative abundance or 0.16 ± 0.62 strains, across all time points and non-intestinal maternal sites combined), mostly found during the first week of life (day 0-7: 2.56% ± 11.4 or 6.1 ± 5.2 strains for non-intestinal sites), and, during that time, involved shared strains from the vaginal (5.13% ± 18.5), skin (3.42% ± 8.24) and oral (0.03% ± 0.15) microbiota of the mother. Thus, ‘vaginal seeding’ of the neonatal microbiota at birth (Mueller et al., 2020) might be less important for the long-term microbiota development of the child than transfer of the intestinal microbiota of the mother.

**Fig. 5:**
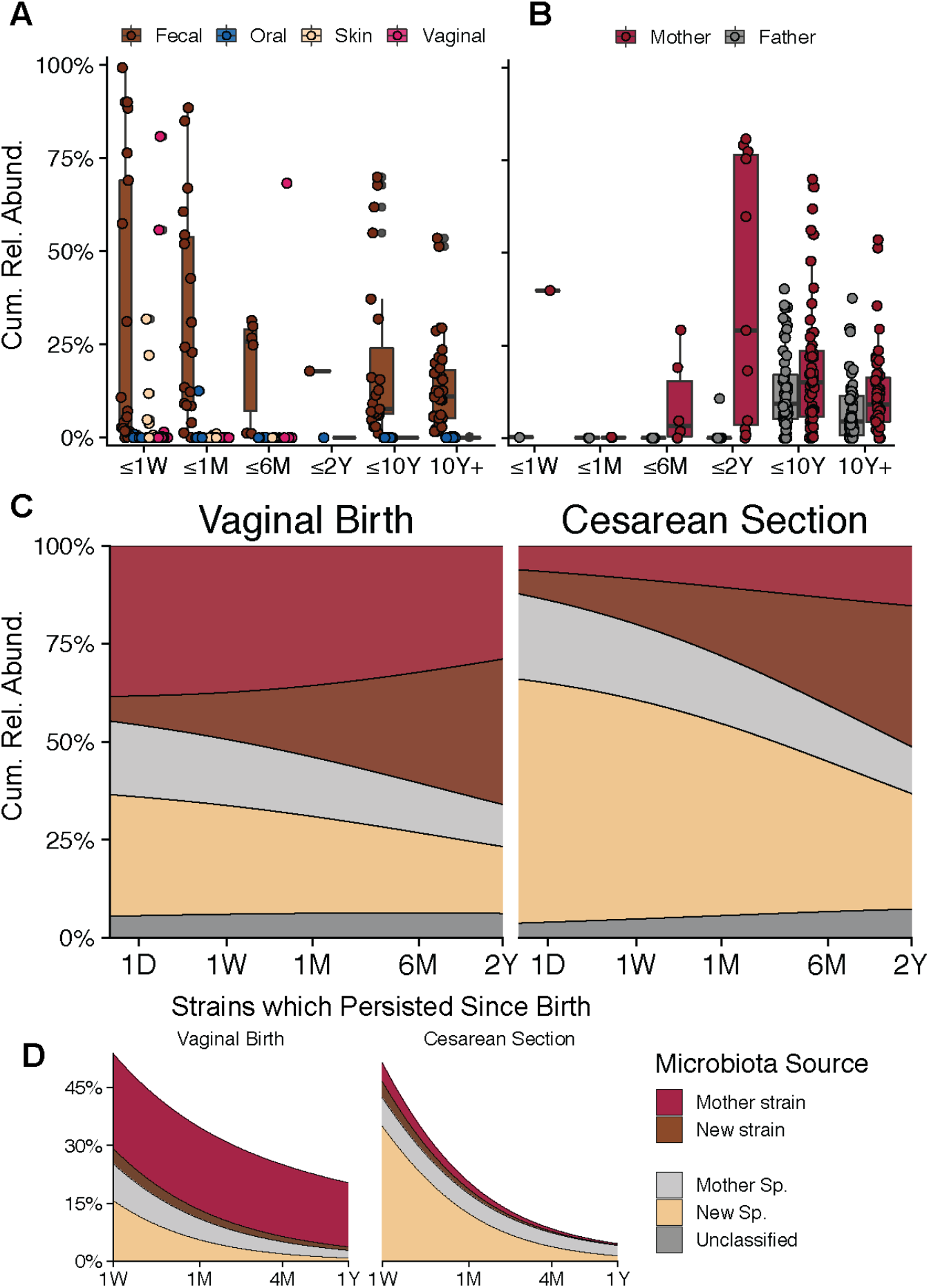
Maternal strain transfer in neonates and persistence through infancy and adolescence. A) Cumulative relative abundances from shared strains between vaginally delivered infants and their mothers mostly involve the maternal fecal microbiota. B) Microbiota fractions from shared strains with the fecal microbiota of the mother and father are increased in older children and adolescents (vaginally born and C-section infants), based on the analysis of available samples from infant-mother-father triads. C) Larger microbiota fractions are represented by maternally shared strains and species in vaginally born compared to C-section infants. See Figure S3 for shared strain counts corresponding to A) B), and C). D) Neonatal microbiota fractions from maternally transferred strains and species (shared at first sampled time point after birth) persist throughout the first year of life. A larger fraction of the first sampled newborn microbiota is lost during the first year of life in C-section compared to vaginally delivered infants.

To also investigate the role of microbial strain transfer from fathers to infants, we used a subset of our combined dataset for which intestinal metagenomic sequence data was available from both parents (Table S1). We determined the maternal and paternal contributions to the child microbiota by quantifying their shared strains and the cumulative relative abundances of their respective species. During the first two years of life, only maternally shared strains contributed substantial fractions to the infant microbiota (Figure 5B; 24.89% ± 30.87 relative abundance or 1.81 ± 1.63 strains vs. 0.52% ± 2.32 relative abundance or 0.29 ± 0.72 strains for fathers). These maternal microbiota contributions remained relatively stable during later infancy, adolescence, and even adulthood (9.5% ± 6.94 relative abundance or 4.59 ± 3.99 strains at >20 years of age), suggesting early transfer of intestinal microbes from the mother to the child, as well as continuous strain persistence, retransfer or mutual exchange between children and their mothers during infancy, adolescence and adulthood. Interestingly, strains that were shared with the intestinal microbiota of the father constituted larger fractions (7.25% ± 8.4 relative abundance or 3.97 ± 3.61 strains) of the older infant, adolescent and adult microbiota (Figure 5B), indicating that microbiota transfer and/or exchange between children and both of their parents continues long after birth and early infancy. Differentiation of infants based on birth type suggests that, during the first year of life, C-section infants harbor reduced numbers of shared strains with their mothers compared to vaginally born infants, but that they compensate for this reduction by sharing more strains with their mothers and fathers later in life (Figure S3B), although the small number of independent samples did not allow for statistical comparisons. The comparison of vaginally delivered and C-section infants with respect to shared strains with the intestinal microbiota from the mother alone (Figure 5C) was statistically better supported (985 infant/mother pairs): Vaginally born infants harbored larger maternally shared microbiota fractions immediately after birth, which remained relatively stable during infancy (day 0-7: 36.56% ± 36.9; ≤2 years: 33.17% ± 25.37; year 2-10: 14.24% ± 9.1). In infants born by C-section, shared strain microbiota contributions were reduced after birth, but increased during infancy and reached levels that were comparable to those from vaginally born infants in childhood (day 0-7: 8.49% ± 21.76; year 2-10: 17.1% ± 17.7). C-section therefore appears to delay rather than abolish maternal strain transfer to the child, at least with respect to the relative abundance fractions of shared strains in children.

Finally, to better understand the dynamics of postpartum microbiota developments in the neonate, we assessed the fate of maternally shared species and strains during the first year of life (Figure 5D). We first identified maternally shared strains in the first available infant sample (collected 0-7 days after birth) and then tracked strain persistence, replacement or coexistence with other strains in available follow-up samples from the same infant. About 75% of the species that were identified immediately after birth were undetectable as early as seven days after birth, representing 50% of the total microbiota at that time (Figure 5D). Thus, the majority of species that colonize the infant gut are not acquired during birth but later in life. However, >10% of the initially identified species in vaginally delivered infants could still be detected one year after birth, accounting for >20% of the total microbiota, compared to only <5% of species and relative abundance in C-section infants.

In summary, C-section appears to most profoundly affect the infant microbiota during the first year of life, resulting in reduced numbers and contributions of shared strains to the infant microbiota, as well as the absence of those maternal species and strains during this developmental period that are transferred at birth.

### Taxonomy-dependent differences in maternal strain sharing

In order to determine if maternal strain transfer rates varied between microbial taxa, all strain and species observations in neonatal and infant samples were classified as representing maternal strains (shared species and strain), maternal species (shared species, but unknown strain), new strains (shared species, but new strain) and new species. The frequencies of events from all four categories were compared for different bacterial genera and between vaginally delivered and C-section infants over time (Figure 6, S4). In vaginally delivered infants, bacterial taxa could roughly be assigned to three distinct groups based on their strain sharing frequency: (i) Species from the genera *Alistipes, Bacteroides, Bilophila, Parabacteroides* and *Sutterella* were more frequently represented by maternal than new, previously undetected, strains (29.66% ± 24.2 maternal strains vs. 10.87% ± 11.15 new strains, on average); (ii) species from the genera *Akkermansia, Bifidobacterium, Collinsella* and *Escherichia* were relatively evenly represented by maternal and new strains (20.1% ± 17.85 maternal strains vs. 26.08% ± 22.08 new strains); and (iii) species from the genera *Blautia, Eubacterium, Faecalibacterium, Ruminococcus* and *Streptococcus* were more frequently represented by new than maternal strains (1.76% ± 3.1 maternal strains vs. 23.88% ± 21.33 new strains). We found at least a partial taxonomic overlap between bacterial strains that were most frequently shared between infants and their mothers and those that were most often shared between adolescents (10 years and older) and their mothers, which most frequently belonged to the genera *Bacteroides, Eubacterium, Paracteroides* and *Sutterella*, suggesting similar taxon-specific patterns of maternal strain sharing in newborns and adult children.

**Fig. 6:**
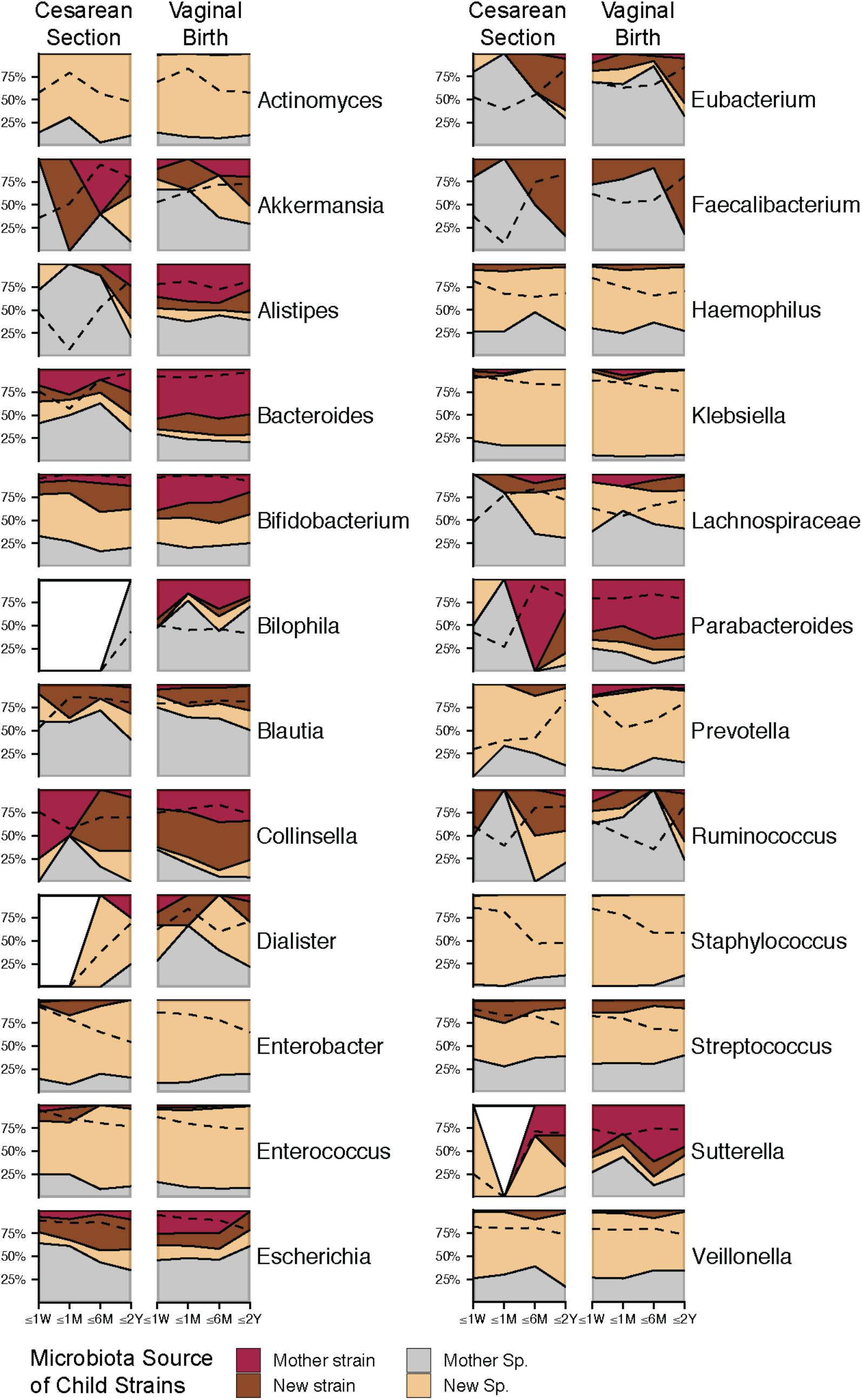
Taxonomy-dependent differences in maternal strain sharing. Each plot shows the fraction of species observations per infant/child sample. Species detected in infants were labeled as “Mother strain” (same species and strain as detected in the mother), “Mother Sp.” (same species as detected in the mother, but strain not resolved), “New strain” (same species as detected in the mother, but different strain), or “New Sp.” (species not detected in the mother). Data are shown as mean frequencies across species of selected genera. Dashed lines indicate genus log-transformed mean relative abundances. Fractions of species observations that were absent in infants are colored white.

In general, sharing of maternal strains and species was observed for the same bacterial taxa in vaginally delivered and C-section infants, but less frequently in C-section infants (Figure 6, S4). Vaginally delivered infants also exhibited less variation in the frequency of maternally shared or new strains over time, which could be partially explained by later maternal microbiota sharing in C-section infants (Figure 5). The genus *Bacteroides* is noteworthy, as it is frequently and consistently represented by maternally shared strains in vaginally delivered infants. Although *Bacteroides* strains are more frequently shared with mothers in C-section infants at >6 months of age, a reduced frequency of shared strains from this genus appears to persist in C-section infants, indicating that potential deficits in microbiota establishment and long-term development should be further investigated at the level of individual bacterial taxa.

## DISCUSSION

The risk for inflammatory and metabolic diseases can be affected by inter-individual taxonomic and functional microbiota variations that result from events at birth and during early infancy, with potential life-long consequences for immune and energy homeostasis (Dominguez-Bello et al., 2019; Torow and Hornef, 2017). Our high-resolution taxonomic meta-analysis of available metagenomics shotgun sequence data presents a detailed and comprehensive picture of the highly dynamic neonatal and infant gut microbiota development. We showed that the developing microbiota continuously and consistently changed over time, as demonstrated before (Stewart et al., 2018; Subramanian et al., 2014), allowing for the development of microbiota-dependent age prediction models. In contrast to previous reports (Bäckhed et al., 2015), C-section was not associated with delayed microbiota maturation. Rather microbiota changes in vaginally born and C-section infants progressed on separate but converging paths, as the variance in taxonomic microbiota compositions between infants that could be explained by birth type decreased over time. Physiologically, these compositional differences resulted in a delayed transition from a predominantly oxygen-tolerant to intolerant microbiota in C-section compared to vaginally born infants. In human adults, differences in luminal intestinal oxygen contents have been associated with dysbiotic microbiota states as a consequence of antibiotic, infectious and inflammatory perturbations (Litvak et al., 2018). In neonatal mouse models of late onset sepsis, decreasing intestinal oxygen levels were linked to the transition from a dysbiotic, facultative anaerobe-dominated to a strict anaerobe-dominated, protective microbiota (Singer et al., 2019). However, in the adult intestine low luminal oxygen is controlled by a positive feedback loop between commensal obligate anaerobes and epithelial cells (Byndloss and Bäumler, 2018). But these pathways appear not to be operational in neonates, as obligate anaerobes could not be engrafted in neonatal mice following fecal microbiota transplantation experiments (Singer et al., 2019). The succession of strict anaerobic bacteria in the neonatal gut has been suggested to result from the consumption of intestinal oxygen by aerobic and facultative anaerobic early colonizers (Favier et al., 2002), which would be consistent with the increased abundance of oxygen-tolerant bacteria that was observed at the intestinal mucosa in mice (Albenberg et al., 2014). As opposed to microbe-mediated mechanisms, a reduction of luminal oxygen has also been attributed to host-dependent control over the neonatal microbiota transition from early colonizing facultative anaerobes to the obligate anaerobes that dominate the infant microbiota and are maintained through adult life (Robertson et al., 2019). Thus, although the specific interplay of microbe and host-mediated intestinal oxygen control remains unclear, the delayed transition from a facultative to strict anaerobe-dominated microbiota in C-section compared to vaginally delivered infants could play a role for risks associated with C-section (Sandall et al., 2018) and should be further studied.

Taxonomic microbiota profiling at subspecies levels provides the opportunity to detect unique shared lineages or strains as evidence for microbial transfer. In this case, the shared strains from a related sample pair (e.g. from corresponding mother-infant dyads) need to be phylogenetically more similar than those from any unrelated sample pair (e.g. from mothers or infants without microbiota contact). To identify individual microbial strains, detect overlapping microbiota strain profiles and map microbial exchange based on unique shared strains, the recently introduced SameStr program from our group was used (Podlesny and Fricke, 2020). As an adaptation of the StrainPhlAn tool (Truong et al., 2017), it uses the clade-specific marker gene database from MetaPhlAn2 (Segata et al., 2012) to detect species-specific strains while also allowing for shared strain calls between non-dominant members of multi-strain species populations - a feature which is not available through StrainPhlAn and increases the rate of detected shared strain events between related but not unrelated samples (Podlesny and Fricke, 2020). The importance of distinguishing individual strains among multi-strain species populations for the characterization of the neonatal microbiota has recently been demonstrated by Yassour et al., as colonization of neonates with dominant and secondary maternal strains from *Bifidobacterium* and *Bacteroides* species was shown to depend on selective advantages based on strain-specific carbohydrate-degrading capabilities (Yassour et al., 2018). Application of SameStr to our combined metagenome dataset resulted in >75000 (3.3% of compared species alignments) detected shared strain events, providing unprecedented statistical support and resolution to characterize microbial exchange between infants and their family members over extended periods of time from immediately after birth to infancy and childhood and into adolescence and adulthood. The specificity and sensitivity of SameStr was showcased with the visualization of shared strain profiles that connected members of the same but not distinct families into family-specific networks. For the first time, these networks also revealed the degree of microbial strain sharing that not only connects mothers and neonates, but also adult children and their mothers, as well as other family members, including infants and their siblings, fathers and grandparents. Together our findings reveal a complex, and so far largely unexplored network of microbiota exchange among family members, illustrating the previously reported dominance of environmental over host genetic influences on human gut microbiota composition and development (Rothschild et al., 2018).

Early infant microbiota studies proposed colonization with the vaginal microbiota of the mother during birth as one of the first and most important events for microbiota initialization (Dominguez-Bello et al., 2010). In the absence of vaginal microbiota exposure, this creates the unfortunate opportunity for other, foreign and potentially disadvantageous microbes to seize upon this ecological niche. These other bacteria have been identified as members of the skin (Dominguez-Bello et al., 2010) or hospital environment microbiota (Shin et al., 2015), including many opportunistic pathogens (Shao et al., 2019). We detected in our meta-analysis only small and transient neonatal gut microbiota fractions that could be attributed to the vaginal microbiota of the mother, either based on functional species classification or the detection of shared species and strains with vaginal metagenomes. The absence of specific gut bacteria that were present in vaginally delivered infants, such as *Bacteroides* and *Bifidobacterium* spp., correlated with increased relative abundance of predicted oral and skin commensals, such as streptococci and staphylococci, in infants born by C-section. Shared strains between infants and their mothers, indicative of maternal transfer, contributed substantial, stable and long-lasting microbiota fractions to the newborn, infant, adolescent and adult microbiota. The intestinal rather than vaginal, skin or oral microbiota of the mother was the most important source of transferred microbes, at least at >2 weeks of age, confirming recent related reports (Ferretti et al., 2018), and this transfer was drastically reduced in infants born by C-section. Reconstitution of the neonatal microbiota by fecal microbiota transplantation (FMT) would therefore appear to be the more obvious therapeutic approach to compensate for the lack of maternal microbiota transfer in C-section infants, as opposed to ‘vaginal seeding’ (Dominguez-Bello et al., 2016). In fact, a recent clinical trial on the use of maternal FMT in infants born by C-section found no adverse effects but increased relative abundance of *Bacteroidaceae* and *Bifidobacteriaceae* compared to untreated infants (Korpela et al., 2020).

The newborn gut microbiota of vaginally delivered infants from our combined dataset included greater contributions of maternally transferred strains than the microbiota of C-section infants. These maternally transferred strains largely persisted through childhood and belonged to the same taxa (*Bacteroides, Parabacteroides*, and *Sutterella*) as bacterial strains that were most often shared between adolescent children and their parents, suggesting potential life-long colonization with maternally transferred strains from these species. However, we also commonly observed strain sharing in older children, in particular at >2 years of age, irrespective of birth type, and including mothers and fathers. Thus, additional mechanisms besides vaginal birth appear to be at play for familial microbiota exchange, which could compensate for the lack of maternal transfer in C-section infants. Detailed phylogenetic analyses of maternally transferred species could be applied to characterize their prevalence, distribution and relationship in different human populations. Similar analyses have been carried out for other members of the human microbiota and could help determine whether the vertical transmission of the species follows a maternal heritability pattern, which could be disrupted in infants born by C-section or replaced with paternal inheritance. Sonnenburg et al. found members of the order *Bacteroidales* to be disproportionately affected by low-fiber diet-induced microbiota depletion in mice (Sonnenburg et al., 2016) and Vatenen et al. presented evidence for immunoinhibitory properties of lipopolysaccharide (LPS) cell wall components from *Bacteroides* species, which were associated with the development of type 1 diabetes (Vatanen et al., 2016). Together, these findings provide a strong justification for additional work on the strain-level characterization of human microbiota initialization and development, including developmental and clinical consequences.

## CONCLUSIONS

Our meta-analysis presents a detailed and comprehensive picture of the highly dynamic neonatal and infant gut microbiota development and presents evidence for taxonomic and functional compositional differences early in life between infants born naturally or by C-section, which persist into adolescence. Our findings encourage further studies to determine whether the disrupted vertical microbiota transfer and lack of maternal strains and species in C-section infants leads to transient developmental microbiota changes or lifelong structural microbiota deficits due to new, altered or missing microbiota members and associated metabolic or immune functions.

## Data Availability

Accession identifiers referring to the published studies and metagenomic shotgun sequencing data used in the analyses can be found in Table 1 of the manuscript.

## Acknowledgements

D.P. and W.F.F. received funding by the German Research Foundation (DFG, Deutsche Forschungsgemeinschaft) under SPP 1656 (Project no. 316130265).

## Supplemental Material

### Supplemental Figures

**Fig. S1.**
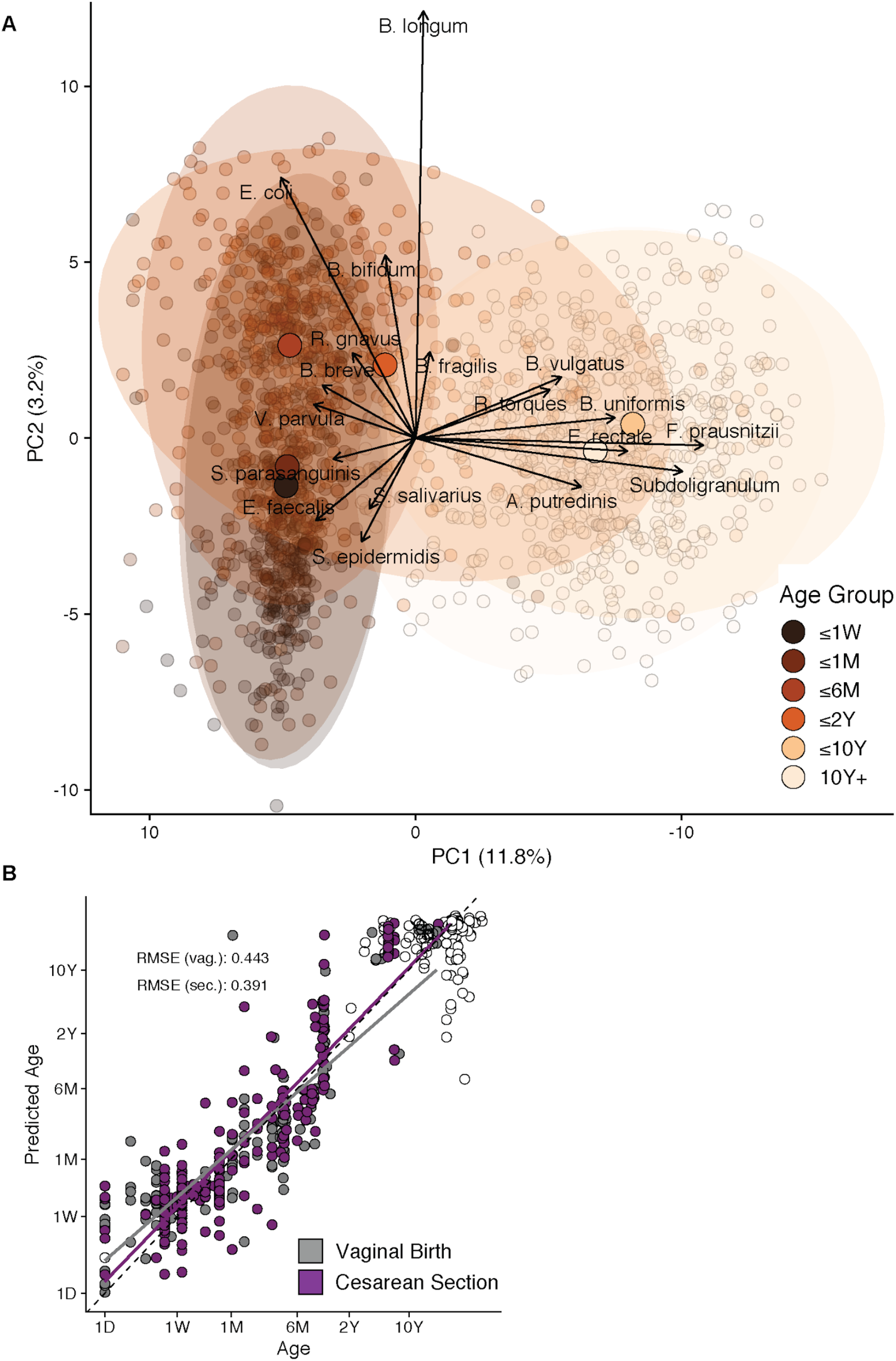
Extends Fig. 1: Maturation of the taxonomic human gut microbiota composition is linked to age. A) Principal component analysis (PCA) biplot of centered log-ratio (clr)-transformed species-level microbial community compositions of human fecal samples obtained from neonatal stage up to late adulthood (range: 0 - 72 years) shows a strong compositional clustering along principal components PC1 and PC2. B) Compositional changes during microbiota maturation are predictive of host age, which is unaffected by the infant delivery mode and does not suggest a general delay in microbiota maturation for infants born by C-section.

**Fig. S2.**
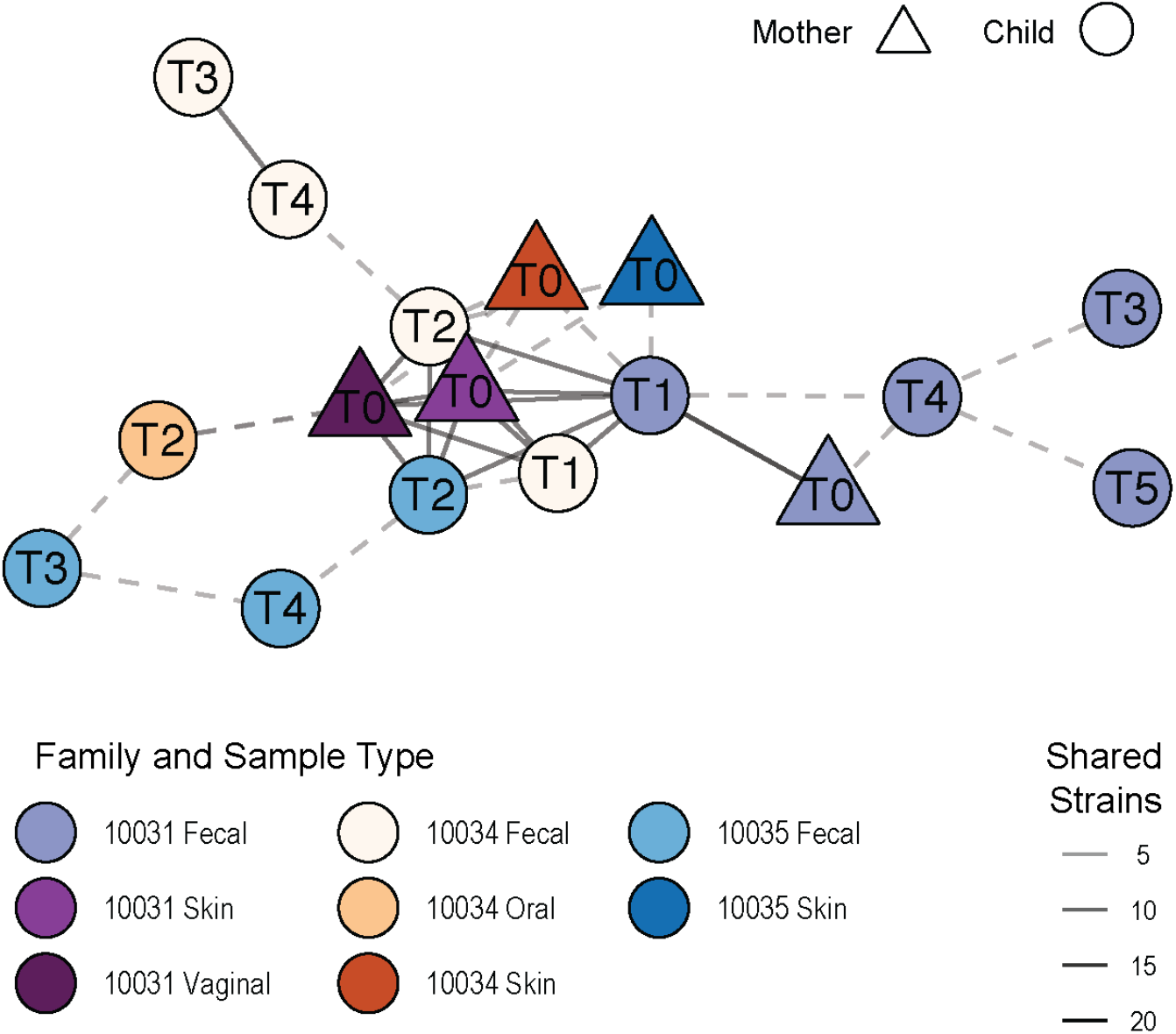
Extends Fig. 4C: Shared strain networks identify microbiota exchange between multi-generational family members and distinct neonates. Network plot showing strain sharing between different samples collected from three newborns that were born at the same neonatal hospital ward and their mothers, numbered according to the sampled day after birth. Five fecal and one oral neonatal samples that were collected during the first two days of life shared strains with each other, as well as with at least one fecal, skin and vaginal samples from each of their mothers. No strain sharing was detected between fecal samples collected at later time points (1 week - 6 months after birth). All three infants shared between 7 and 9 strains with each other. Dashed lines show connections with 2-4 shared strains.

**Fig. S3.**
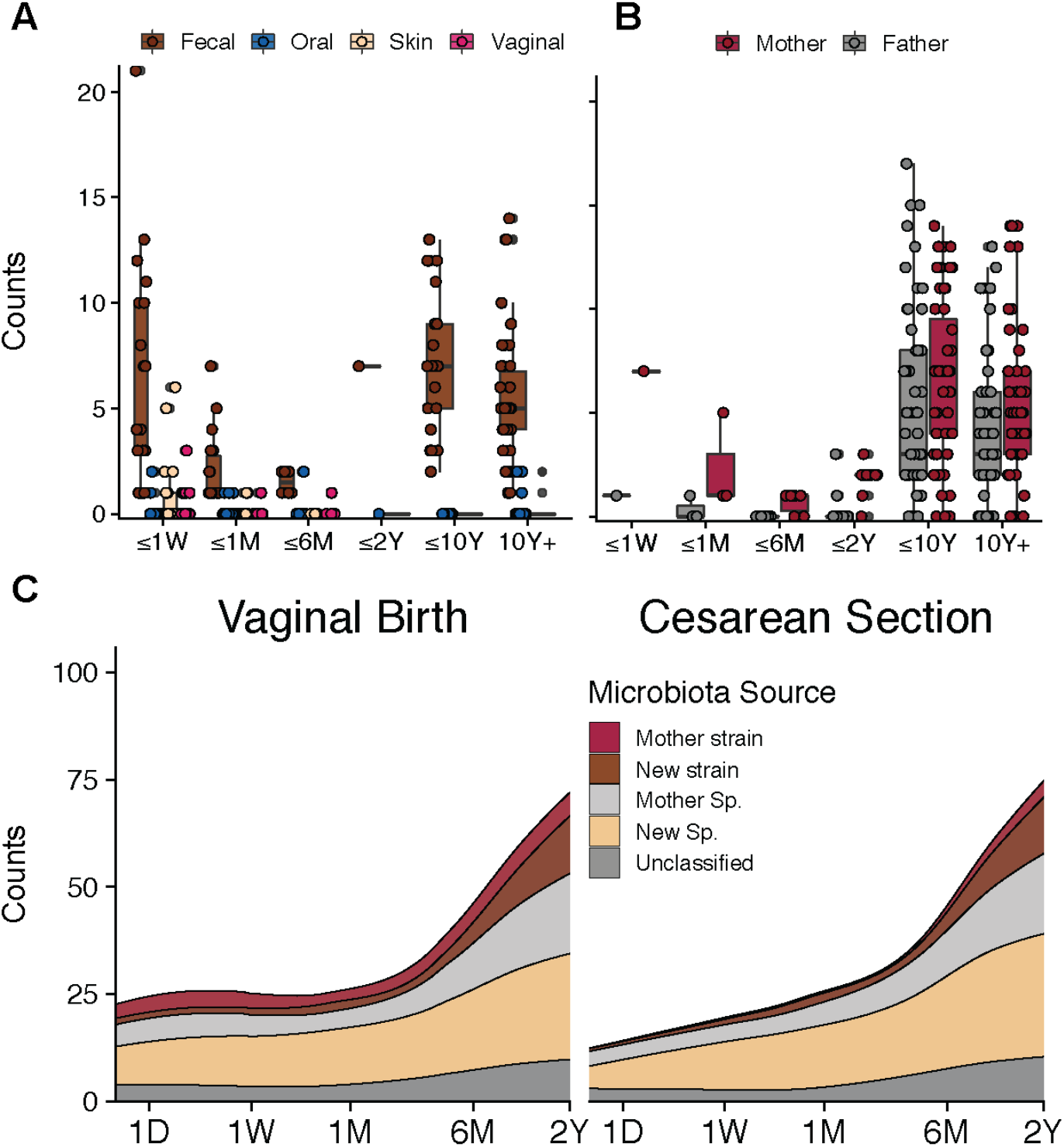
Extends Fig. 5: Maternal strain transfer in neonates and persistence through infancy and adolescence. A) Total counts of shared strains between the infant gut microbiota and metagenomes from different body sites of the mother mostly show strain sharing with the maternal fecal microbiota. B) Numbers of shared strains with the fecal microbiota of the mother and father are increased in older children and adolescents, based on the analysis of available samples from infant-mother-father triads. C) Shared strain numbers between neonates and the fecal microbiota of the mother are higher in vaginally born compared to C-section infants throughout the first two years of life.

**Fig. S4.**
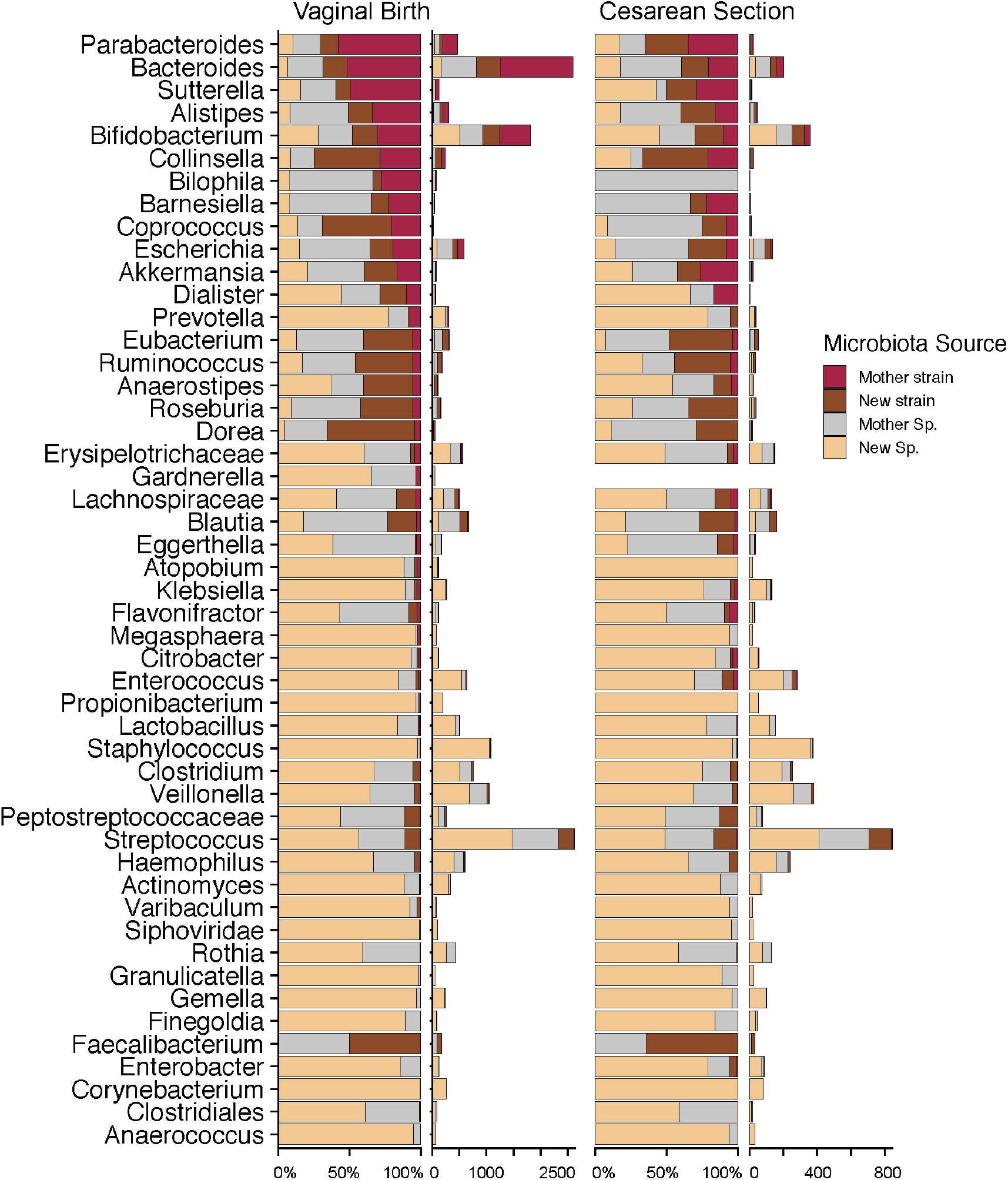
Extends Fig. 6: Taxonomy-dependent differences in maternal strain sharing. Bacterial genus-specific frequencies of strain and species sharing within the first two years of life are shown for infants born by vaginal birth and C-section. For each group, bars show the fraction (left) and the total number (right) of species observations, which were assigned to four categories: “Mother strain” (same species and strain as detected in the mother), “Mother Sp.” (same species as detected in the mother, but strain not resolved), “New strain” (same species as detected in the mother, but different strain), or “New Sp.” (species not detected in the mother). Data are shown as mean frequencies across species of selected genera.

## References

Albenberg, L., Esipova, T.V., Judge, C.P., Bittinger, K., Chen, J., Laughlin, A., Grunberg, S., Baldassano, R.N., Lewis, J.D., Li, H., Thom, S.R., Bushman, F.D., Vinogradov, S.A., Wu, G.D., 2014. Correlation between intraluminal oxygen gradient and radial partitioning of intestinal microbiota. Gastroenterology 147, 1055–63.e8.

Arrieta, M.-C., Stiemsma, L.T., Amenyogbe, N., Brown, E.M., Finlay, B., 2014. The intestinal microbiome in early life: health and disease. Front. Immunol. 5, 427.

Asnicar, F., Manara, S., Zolfo, M., Truong, D.T., Scholz, M., Armanini, F., Ferretti, P., Gorfer, V., Pedrotti, A., Tett, A., Segata, N., 2017. Studying Vertical Microbiome Transmission from Mothers to Infants by Strain-Level Metagenomic Profiling. mSystems 2. https://doi.org/10.1128/mSystems.00164-16

Bäckhed, F., Roswall, J., Peng, Y., Feng, Q., Jia, H., Kovatcheva-Datchary, P., Li, Y., Xia, Y., Xie, H., Zhong, H., Khan, M.T., Zhang, J., Li, J., Xiao, L., Al-Aama, J., Zhang, D., Lee, Y.S., Kotowska, D., Colding, C., Tremaroli, V., Yin, Y., Bergman, S., Xu, X., Madsen, L., Kristiansen, K., Dahlgren, J., Wang, J., 2015. Dynamics and Stabilization of the Human Gut Microbiome during the First Year of Life. Cell Host Microbe 17, 690–703.

Bazanella, M., Maier, T.V., Clavel, T., Lagkouvardos, I., Lucio, M., Maldonado-Gòmez, M.X., Autran, C., Walter, J., Bode, L., Schmitt-Kopplin, P., Haller, D., 2017. Randomized controlled trial on the impact of early-life intervention with bifidobacteria on the healthy infant fecal microbiota and metabolome. Am. J. Clin. Nutr. 106, 1274–1286.

Blaser, M.J., Dominguez-Bello, M.G., 2016. The Human Microbiome before Birth. Cell Host Microbe 20, 558–560.

Bokulich, N.A., Chung, J., Battaglia, T., Henderson, N., Jay, M., Li, H. D Lieber, A., Wu, F., Perez-Perez, G.I., Chen, Y., Schweizer, W., Zheng, X., Contreras, M., Dominguez-Bello, M.G., Blaser, M.J., 2016. Antibiotics, birth mode, and diet shape microbiome maturation during early life. Sci. Transl. Med. 8, 343ra82.

Byndloss, M.X., Bäumler, A.J., 2018. The germ-organ theory of non-communicable diseases. Nat. Rev. Microbiol. 16, 103–110.

Cho, I., Blaser, M.J., 2012. The human microbiome: at the interface of health and disease. Nat. Rev. Genet. 13, 260–270.

Chu, D.M., Ma, J., Prince, A.L., Antony, K.M., Seferovic, M.D., Aagaard, K.M., 2017. Maturation of the infant microbiome community structure and function across multiple body sites and in relation to mode of delivery. Nat. Med. 23, 314–326.

Costello, E.K., Lauber, C.L., Hamady, M., Fierer, N., Gordon, J.I., Knight, R., 2009. Bacterial community variation in human body habitats across space and time. Science 326, 1694–1697.

Cunnington, A.J., Sim, K., Deierl, A., Kroll, J.S., Brannigan, E., Darby, J., 2016. “Vaginal seeding” of infants born by caesarean section. BMJ 352, i227.

Dominguez-Bello, M.G., Costello, E.K., Contreras, M., Magris, M., Hidalgo, G., Fierer, N., Knight, R., 2010. Delivery mode shapes the acquisition and structure of the initial microbiota across multiple body habitats in newborns. Proc. Natl. Acad. Sci. U. S. A. 107, 11971–11975.

Dominguez-Bello, M.G., De Jesus-Laboy, K.M., Shen, N., Cox, L.M., Amir, A., Gonzalez, A., Bokulich, N.A., Song, S.J., Hoashi, M., Rivera-Vinas, J.I., Mendez, K., Knight, R., Clemente, J.C., 2016. Partial restoration of the microbiota of cesarean-born infants via vaginal microbial transfer. Nat. Med. 22, 250–253.

Dominguez-Bello, M.G., Godoy-Vitorino, F., Knight, R., Blaser, M.J., 2019. Role of the microbiome in human development. Gut 68, 1108–1114.

Favier, C.F., Vaughan, E.E., De Vos, W.M., Akkermans, A.D.L., 2002. Molecular monitoring of succession of bacterial communities in human neonates. Appl. Environ. Microbiol. 68, 219–226.

Ferretti, P., Pasolli, E., Tett, A., Asnicar, F., Gorfer, V., Fedi, S., Armanini, F., Truong, D.T., Manara, S., Zolfo, M., Beghini, F., Bertorelli, R., De Sanctis, V., Bariletti, I., Canto, R., Clementi, R., Cologna, M., Crifò, T., Cusumano, G., Gottardi, S., Innamorati, C., Masè, C., Postai, D., Savoi, D., Duranti, S., Lugli, G.A., Mancabelli, L., Turroni, F., Ferrario, C., Milani, C., Mangifesta, M., Anzalone, R., Viappiani, A., Yassour, M., Vlamakis, H., Xavier, R., Collado, C.M., Koren, O., Tateo, S., Soffiati, M., Pedrotti, A., Ventura, M., Huttenhower, C., Bork, P., Segata, N., 2018. Mother-to-Infant Microbial Transmission from Different Body Sites Shapes the Developing Infant Gut Microbiome. Cell Host Microbe 24, 133–145.e5.

Galazzo, G., van Best, N., Bervoets, L., Dapaah, I.O., Savelkoul, P.H., Hornef, M.W., GI-MDH consortium, Lau, S., Hamelmann, E., Penders, J., 2020. Development of the Microbiota and Associations With Birth Mode, Diet, and Atopic Disorders in a Longitudinal Analysis of Stool Samples, Collected From Infancy Through Early Childhood. Gastroenterology 158, 1584–1596.

Heintz-Buschart, A., May, P., Laczny, C.C., Lebrun, L.A., Bellora, C., Krishna, A., Wampach, L., Schneider, J.G., Hogan, A., de Beaufort, C., Wilmes, P., 2016. Integrated multi-omics of the human gut microbiome in a case study of familial type 1 diabetes. Nature Microbiology 2, 1–13.

Hornef, M., Penders, J., 2017. Does a prenatal bacterial microbiota exist? Mucosal Immunol. 10, 598–601.

Korpela, K., Costea, P., Coelho, L.P., Kandels-Lewis, S., Willemsen, G., Boomsma, D.I., Segata, N., Bork, P., 2018. Selective maternal seeding and environment shape the human gut microbiome. Genome Res. 28, 561–568.

Korpela, K., Helve, O., Kolho, K.-L., Saisto, T., Skogberg, K., Dikareva, E., Stefanovic, V., Salonen, A., Andersson, S., de Vos, W.M., 2020. Maternal Fecal Microbiota Transplantation in Cesarean-Born Infants Rapidly Restores Normal Gut Microbial Development: A Proof-of-Concept Study. Cell. https://doi.org/10.1016/j.cell.2020.08.047

Litvak, Y., Byndloss, M.X., Bäumler, A.J., 2018. Colonocyte metabolism shapes the gut microbiota. Science 362. https://doi.org/10.1126/science.aat9076

Lloyd-Price, J., Mahurkar, A., Rahnavard, G., Crabtree, J., Orvis, J., Hall, A.B., Brady, A., Creasy, H.H., McCracken, C., Giglio, M.G., McDonald, D., Franzosa, E.A., Knight, R., White, O., Huttenhower, C., 2017. Strains, functions and dynamics in the expanded Human Microbiome Project. Nature 550, 61–66.

Mueller, N.T., Dominguez-Bello, M.G., Appel, L.J., Hourigan, S.K., 2020. “Vaginal seeding” after a caesarean section provides benefits to newborn children: FOR: Does exposing caesarean-delivered newborns to the vaginal microbiome affect their chronic disease risk? The critical need for trials of “vaginal seeding” during caesarean section. BJOG 127, 301.

Mueller, N.T., Whyatt, R., Hoepner, L., Oberfield, S., Dominguez-Bello, M.G., Widen, E.M., Hassoun, A., Perera, F., Rundle, A., 2015. Prenatal exposure to antibiotics, cesarean section and risk of childhood obesity. Int. J. Obes. 39, 665–670.

Podlesny, D., Fricke, W.F., 2020. Microbial Strain Engraftment, Persistence and Replacement after Fecal Microbiota Transplantation. Preprint in medRxiv. https://doi.org/10.1101/2020.09.29.20203638.

Reyman, M., van Houten, M.A., van Baarle, D., Bosch, A.A.T.M., Man, W.H., Chu, M.L.J.N., Arp, K., Watson, R.L., Sanders, E.A.M., Fuentes, S., Bogaert, D., 2019. Impact of delivery mode-associated gut microbiota dynamics on health in the first year of life. Nat. Commun. 10, 4997.

Robertson, R.C., Manges, A.R., Finlay, B.B., Prendergast, A.J., 2019. The Human Microbiome and Child Growth - First 1000 Days and Beyond. Trends Microbiol. 27, 131–147.

Rothschild, D., Weissbrod, O., Barkan, E., Kurilshikov, A., Korem, T., Zeevi, D., Costea, P.I., Godneva, A., Kalka, I.N., Bar, N., Shilo, S., Lador, D., Vila, A.V., Zmora, N., Pevsner-Fischer, M., Israeli, D., Kosower, N., Malka, G., Wolf, B.C., Avnit-Sagi, T., Lotan-Pompan, M., Weinberger, A., Halpern, Z., Carmi, S., Fu, J., Wijmenga, C., Zhernakova, A., Elinav, E., Segal, E., 2018. Environment dominates over host genetics in shaping human gut microbiota. Nature 555, 210–215.

Sandall, J., Tribe, R.M., Avery, L., Mola, G., Visser, G.H., Homer, C.S., Gibbons, D., Kelly, N.M., Kennedy, H.P., Kidanto, H., Taylor, P., Temmerman, M., 2018. Short-term and long-term effects of caesarean section on the health of women and children. Lancet 392, 1349–1357.

Schloss, P.D., Iverson, K.D., Petrosino, J.F., Schloss, S.J., 2014. The dynamics of a family’s gut microbiota reveal variations on a theme. Microbiome 2, 1–13.

Segata, N., Waldron, L., Ballarini, A., Narasimhan, V., Jousson, O., Huttenhower, C., 2012. Metagenomic microbial community profiling using unique clade-specific marker genes. Nat. Methods 9, 811–814.

Shao, Y., Forster, S.C., Tsaliki, E., Vervier, K., Strang, A., Simpson, N., Kumar, N., Stares, M.D., Rodger, A., Brocklehurst, P., Field, N., Lawley, T.D., 2019. Stunted microbiota and opportunistic pathogen colonization in caesarean-section birth. Nature 574, 117–121.

Shin, H., Pei, Z., Martinez, K.A., 2nd, Rivera-Vinas, J.I., Mendez, K., Cavallin, H., Dominguez-Bello, M.G., 2015. The first microbial environment of infants born by C-section: the operating room microbes. Microbiome 3, 59.

Singer, J.R., Blosser, E.G., Zindl, C.L., Silberger, D.J., Conlan, S., Laufer, V.A., DiToro, D., Deming, C., Kumar, R., Morrow, C.D., Segre, J.A., Gray, M.J., Randolph, D.A., Weaver, C.T., 2019. Preventing dysbiosis of the neonatal mouse intestinal microbiome protects against late-onset sepsis. Nat. Med. 25, 1772–1782.

Sonnenburg, E.D., Smits, S.A., Tikhonov, M., Higginbottom, S.K., Wingreen, N.S., Sonnenburg, J.L., 2016. Diet-induced extinctions in the gut microbiota compound over generations. Nature 529, 212–215.

Stewart, C.J., Ajami, N.J., O’Brien, J.L., Hutchinson, D.S., Smith, D.P., Wong, M.C., Ross, M.C., Lloyd, R.E., Doddapaneni, H., Metcalf, G.A., Muzny, D., Gibbs, R.A., Vatanen, T., Huttenhower, C., Xavier, R.J., Rewers, M., Hagopian, W., Toppari, J., Ziegler, A.-G., She, J.-X., Akolkar, B., Lernmark, A., Hyoty, H., Vehik, K., Krischer, J.P., Petrosino, J.F., 2018. Temporal development of the gut microbiome in early childhood from the TEDDY study. Nature 562, 583–588.

Stinson, L.F., Payne, M.S., Keelan, J.A., 2018. A Critical Review of the Bacterial Baptism Hypothesis and the Impact of Cesarean Delivery on the Infant Microbiome. Front. Med. 5, 135.

Subramanian, S., Huq, S., Yatsunenko, T., Haque, R., Mahfuz, M., Alam, M.A., Benezra, A., DeStefano, J., Meier, M.F., Muegge, B.D., Barratt, M.J., VanArendonk, L.G., Zhang, Q., Province, M.A., Petri, W.A., Jr, Ahmed, T., Gordon, J.I., 2014. Persistent gut microbiota immaturity in malnourished Bangladeshi children. Nature 510, 417–421.

Torow, N., Hornef, M.W., 2017. The Neonatal Window of Opportunity: Setting the Stage for Life-Long Host-Microbial Interaction and Immune Homeostasis. J. Immunol. 198, 557–563.

Truong, D.T., Franzosa, E.A., Tickle, T.L., Scholz, M., Weingart, G., Pasolli, E., Tett, A., Huttenhower, C., Segata, N., 2015. MetaPhlAn2 for enhanced metagenomic taxonomic profiling. Nat. Methods 12, 902–903.

Truong, D.T., Tett, A., Pasolli, E., Huttenhower, C., Segata, N., 2017. Microbial strain-level population structure and genetic diversity from metagenomes. Genome Res. 27, 626–638.

Van Rossum, T., Ferretti, P., Maistrenko, O.M., Bork, P., 2020. Diversity within species: interpreting strains in microbiomes. Nat. Rev. Microbiol. 18, 491–506.

Vatanen, T., Kostic, A.D., d’Hennezel, E., Siljander, H., Franzosa, E.A., Yassour, M., Kolde, R., Vlamakis, H., Arthur, T.D., Hämäläinen, A.-M., Peet, A., Tillmann, V., Uibo, R., Mokurov, S., Dorshakova, N., Ilonen, J., Virtanen, S.M., Szabo, S.J., Porter, J.A., Lähdesmäki, H., Huttenhower, C., Gevers, D., Cullen, T.W., Knip, M., DIABIMMUNE Study Group, Xavier, R.J., 2016. Variation in Microbiome LPS Immunogenicity Contributes to Autoimmunity in Humans. Cell 165, 842–853.

Yan, Y., Nguyen, L.H., Franzosa, E.A., Huttenhower, C., 2020. Strain-level epidemiology of microbial communities and the human microbiome. Genome Med. 12, 71.

Yassour, M., Jason, E., Hogstrom, L.J., Arthur, T.D., Tripathi, S., Siljander, H., Selvenius, J., Oikarinen, S., Hyöty, H., Virtanen, S.M., Ilonen, J., Ferretti, P., Pasolli, E., Tett, A., Asnicar, F., Segata, N., Vlamakis, H., Lander, E.S., Huttenhower, C., Knip, M., Xavier, R.J., 2018. Strain-Level Analysis of Mother-to-Child Bacterial Transmission during the First Few Months of Life. Cell Host Microbe 24, 146–154.e4.

Zuo, T., Kamm, M.A., Colombel, J.-F., Ng, S.C., 2018. Urbanization and the gut microbiota in health and inflammatory bowel disease. Nat. Rev. Gastroenterol. Hepatol. 15, 440–452.

